# Visit-to-Visit Blood Pressure Variability, Glycemic Status and the Risk of Dementia, Stroke, and All-Cause Mortality: A UK Biobank Cohort Study

**DOI:** 10.64898/2026.10.11.26365219

**Authors:** Si Han, Jesper de Jong, Yingying Zhou, Miriam Sturkenboom, Geert Jan Biessels, Fariba Ahmadizar

## Abstract

**Aims:** The risks of dementia, stroke, and mortality increase from normoglycemia to prediabetes and type 2 diabetes (T2D). Blood pressure variability (BPV) is associated with adverse vascular and neurological outcomes, but its role in glycemia-associated risks remains unclear. We examined the interplay between glycemia, BPV, and these outcomes.

**Methods and Results:** In a large prospective cohort, visit-to-visit BPV was quantified using absolute and directional measures of systolic (SBP), diastolic (DBP), mean arterial (MAP), and pulse pressure (PP). Glycemic status was defined as normoglycemia, prediabetes, and T2D. Time-updated Cox models estimated outcome associations, adjusting for confounders and mean blood pressure. We evaluated the joint and independent associations of BPV and glycemic status with outcomes, effect modification, and attenuation of glycemia-associated risks after BPV adjustment.

Among 57,611 participants (median follow-up, 5.1 years; IQR, 3.3-9.5 years), compared with normoglycemia and low BPV, T2D with high SBP variability was associated with higher risks of all-cause mortality (HR 2.80, 95% CI 2.12-3.69) and nominally higher risk of all-cause dementia (HR 2.46, 95% CI 1.37-4.42). All absolute BPV measures were independently associated with mortality, while DBP and MAP variability were associated with overall stroke after FDR correction. Additional adjustment for BPV minimally attenuated glycemia-associated risk, and no multiplicative interaction remained significant after multiple-testing correction.

**Conclusions:** Absolute BPV was associated with mortality, and DBP and MAP variability with overall stroke, after accounting for glycemic status and mean blood pressure. Additional BPV adjustment changed the glycemic association only minimally.

## Introduction

Type 2 diabetes (T2D) and prediabetes are increasingly prevalent worldwide and represent major public health challenges. More than 500 million adults are estimated to be living with diabetes globally, and this number is projected to rise to 900 million by 2050 (1, 2). Beyond their well-established metabolic consequences, both prediabetes and T2D are associated with increased risks of stroke, dementia, and premature mortality (3–5). Notably, excess risk appears to emerge before overt diabetes, suggesting that vascular and metabolic dysfunction accumulate progressively across the glycemic spectrum (5, 6). However, the mechanisms underlying these associations remain incompletely understood.

Blood pressure variability (BPV), defined as fluctuations in blood pressure over time, has emerged as a potential independent marker of cardiovascular risk (7, 8). Growing evidence suggests that elevated visit-to-visit BPV is associated with increased risks of stroke, dementia, and all-cause mortality, independent of mean blood pressure, and these associations persist after accounting for antihypertensive treatment (9–16). Proposed mechanisms include impaired cerebral autoregulation, increased arterial stiffness, endothelial dysfunction, and cumulative vascular injury, all of which may contribute to cerebrovascular disease and dementia (12, 17).

Because individuals with prediabetes and T2D already have underlying vascular and autonomic dysfunction, they may be particularly susceptible to these adverse effects of BPV. However, it remains unclear whether BPV provides prognostic information beyond glycemic status, whether individuals with both dysglycemia and elevated BPV have particularly high risks, and whether the associations between BPV and adverse outcomes differ across the glycemic spectrum.

Therefore, in this large prospective cohort study with repeated blood pressure measurements, we investigated the relationships between glycemic states and the risks of dementia (including all-cause dementia, Alzheimer’s disease [AD], and vascular dementia [VaD]), stroke (including ischemic [IS] and hemorrhagic [ICH] subtypes), and all-cause mortality, and especially assessed whether these associations were modified by BPV. Specifically, we aimed to (1) evaluate whether BPV provides independent and complementary prognostic information beyond glycemic status, (2) evaluate the joint associations of glycemic status and BPV with these outcomes, and (3) assess whether BPV modifies the associations between glycemic status and adverse outcomes.

## Methods

### 2.1 Study Setting

The UK Biobank (UKB) is a large prospective population-based cohort that recruited more than 500,000 adults aged 40-69 years between 2006 and 2010. At baseline, participants completed touchscreen questionnaires and interviewer-administered assessments, underwent standardized physical measurements, and provided biological samples at one of 22 assessment centers across England, Scotland, and Wales. The study was approved by the Northwest Multicenter Research Ethics Committee (reference 16/NW/0274), and all participants provided written informed consent.

The primary exposure was baseline glycemic status, classified as normoglycemia (glycated hemoglobin [HbA1c] <5.7% [<39 mmol/mol]), prediabetes (HbA1c 5.7-6.4% [39-47 mmol/mol] without glucose-lowering medication), or T2D. We identified T2D using an established algorithm integrating self-reported physician diagnosis, glucose-lowering medication use, hospital inpatient records, and biochemical measurements, as described previously (18, 19). Hospital-recorded T2D was defined using ICD-9 codes 250.00, 250.10, 250.20, and 250.90 or ICD-10 code E11. The classification algorithm is presented in *Supplementary Figure S1*.

### 2.2 Study Population

Participants were eligible for inclusion if they had valid blood pressure measurements at baseline (visit 0) and at least one subsequent assessment visit with complete BP data (visit 1, 2, or 3), allowing construction of at least one BPV interval *(Figure 1)*. Because BPV was defined using pairs of consecutive visits, participants did not enter the risk set until the later visit used in the first BPV calculation. Accordingly, follow-up began at the first assessment visit after baseline with complete BP data. For a visit to be considered complete, two systolic blood pressure (SBP) and two diastolic blood pressure (DBP) measurements had to be available, obtained either via automated readings or, where unavailable, via manual readings. Automatic measurements were prioritized over manual measurements when both were present. Participants without valid baseline BP or without at least one follow-up BP measurement were excluded. To reduce the influence of implausible values, extreme outliers (>5 standard deviations from the mean) were excluded for continuous variables. A complete-case approach was used, as the proportion of missing data was low across covariates; complete-case analysis was considered unlikely to materially affect the findings.

**Figure 1.**
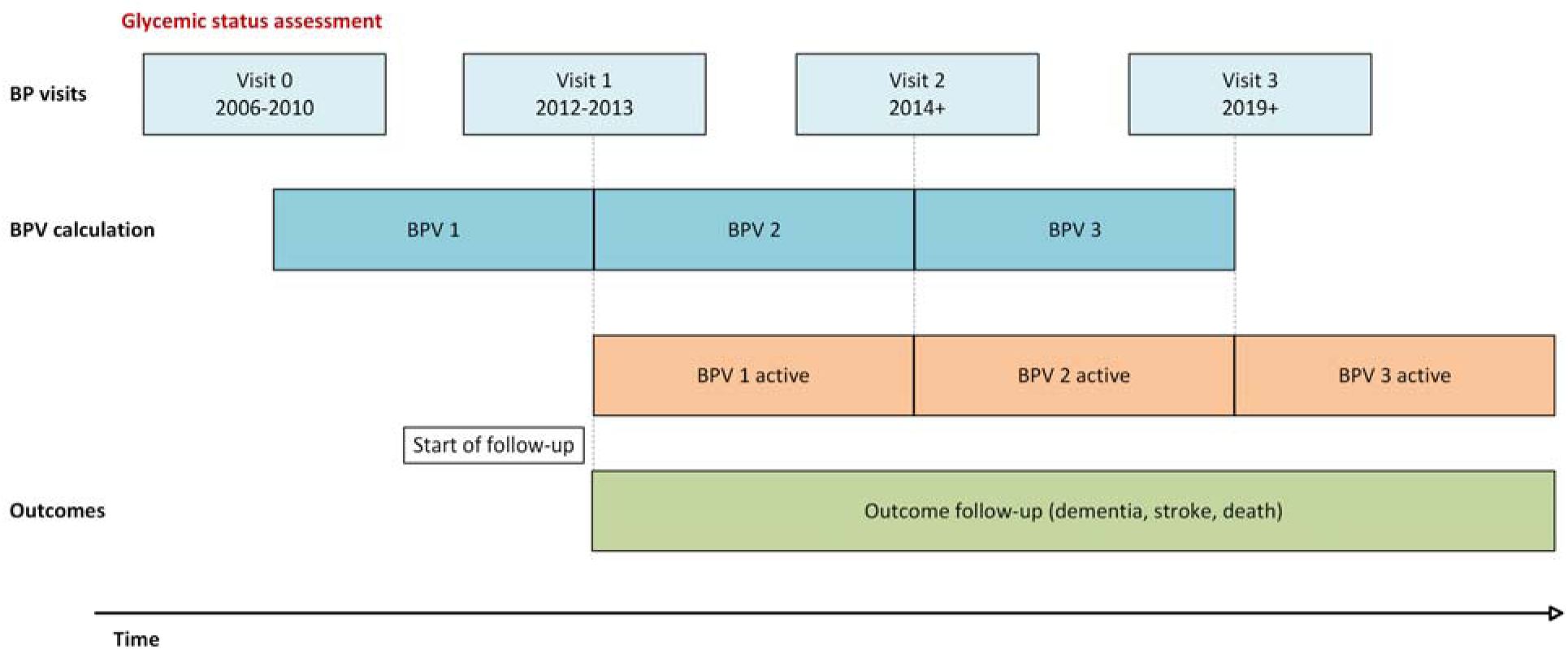
Study design illustrating construction of time-updated blood pressure variability (BPV) and analytical follow-up **Abbreviations:** BP, blood pressure; BPV, blood pressure variability. **Notes:** Glycemic status was assessed at Visit 0. BPV was calculated between consecutive available visits with complete BP measurements. When an intermediate visit was missing, the next available visit was used (e.g., Visit 0–2 or Visit 0–3). Participants entered the risk set at their first eligible repeat BP assessment using delayed entry (left truncation). During follow-up, BPV was treated as a time-varying exposure, with each estimate becoming active at the later visit and remaining in effect until the next BP assessment, outcome event, loss to follow-up, or administrative censoring. **Alt text:** Timeline illustrating the study design and construction of time-updated blood pressure variability. Glycemic status was assessed at Visit 0. Blood pressure variability was calculated between consecutive available visits with complete blood pressure measurements. Follow-up began at the second eligible blood pressure assessment, with each variability estimate becoming active at the later visit used for its calculation and remaining in effect until the next assessment or censoring. Participants were followed for dementia, stroke, and death.

### 2.3 Blood Pressure Assessment

Blood pressure was measured at UKB assessment centers using standardized protocols. At each visit, SBP and DBP were recorded as the mean of two measurements. Mean arterial pressure (MAP) and pulse pressure (PP) were derived for each visit as:

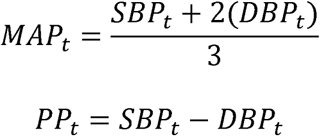

Where *t* denotes the assessment visit.

Visit-to-visit BPV was calculated between consecutive available BP assessments across up to four visits (Visits 0-3). Participants were required to have valid BP measurements at baseline and at least one follow-up visit. If an intermediate visit was missing, the next available assessment was used (e.g., Visits 0-2 or 0-3). Follow-up began at the first eligible repeat visit, and BPV was updated as additional measurements became available. For each BP measure (SBP, DBP, MAP, and PP), two BPV metrics were calculated: absolute variability, defined as the absolute difference between consecutive measurements divided by their mean, and directional variability, defined as the signed difference between consecutive measurements divided by their mean:

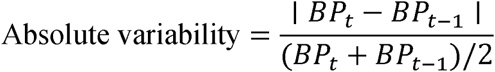

To account for unequal intervals between visits, all BPV measures were annualized by dividing by the time between visits (in years).

For each visit pair, the corresponding interval mean BP (SBP, DBP, MAP, or PP) was calculated as the average of the current and previous measurements and included in the models to distinguish the effect of BP variability from those of average BP level. BPV and interval mean BP were modelled as time-varying exposures. Each new BPV value became active at the later of the two visits used to calculate it and remained in effect until the next BP assessment or the end of follow-up. For participants with multiple repeated assessments, follow-up time was split into consecutive observation intervals defined by adjacent assessment visits. BPV and interval mean BP calculated from a given visit pair were assigned to the interval beginning at the later visit and carried forward until the next available BP assessment, event occurrence, death, or censoring.

BPV measures were standardized as z-scores using the cohort-specific mean and standard deviation to facilitate direct comparison across BPV metrics, including both absolute and directional measures:

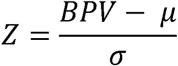

where μ and σ denote the mean and standard deviation of the corresponding BPV measure, respectively. Effect estimates are presented per 1 standard deviation (SD) increase in BPV. Spearman correlation coefficients were calculated to assess the correlations among the BPV measures (SBPV, DBPV, MAPV, and PPV). For analyses requiring categorical classification, BPV was additionally categorized into tertiles and represented low, intermediate, and high BPV, with the lowest tertile serving as the reference category.

### 2.4 Outcome Ascertainment

Follow-up began at the start-of-follow-up date, defined as the first visit with complete BP data after baseline assessment. This corresponded to the second visit used in the first BPV calculation. Follow-up ended at the earliest of the incident event of interest, death, or administrative censoring. The study outcomes were incident all-cause dementia, AD, VaD, stroke, IS, ICH, and all-cause mortality. Outcome dates were derived from linked electronic health records and registry sources. For each outcome, the earliest recorded diagnosis date across available data sources was used. We defined all-cause mortality as death from any cause during follow-up. The date of death was obtained from the death registry. Participants who experienced the outcome of interest before the first eligible BPV assessment interval were not included in the corresponding risk set, because BPV had not yet been defined. Because BPV requires at least two BP measurements, the start of follow-up for the survival analyses was defined as the first subsequent visit at which BPV could be calculated. Thus, the cohort baseline and the analytical start of follow-up were distinct.

### 2.5 Covariate Assessment

Baseline covariates were assessed at the UKB baseline visit. Covariates were selected based on clinical relevance and data availability. These included baseline age, sex, ethnic group, education, Townsend index, body mass index (BMI), smoking status, alcohol consumption, use of blood pressure-lowering medication, use of lipid-lowering medication, and history of vascular disease. Sex was included as a binary variable, and ethnic background was coded according to UKB categories. BMI was modeled as a continuous variable, and smoking status as a categorical variable. History of vascular disease was defined using hospital diagnosis codes (ICD-10 codes I20-I25, I60-I69, and I70-I79). In addition, interval mean blood pressure was included in all models to account for average BP level and to isolate the independent effect of BP variability.

### 2.6 Statistical Analysis

Baseline characteristics were summarized by glycemic status as mean (SD) for approximately normally distributed continuous variables, median (interquartile range [IQR]) for skewed continuous variables, and n (%) for categorical variables. Percentages were calculated among participants with non-missing data for each variable. Differences across glycemic-status groups were assessed using Analysis of variance (ANOVA) for approximately normally distributed continuous variables, the Kruskal-Wallis test for skewed continuous variables, and Fisher’s exact test for categorical variables.

For the analyses, first, independent associations between glycemic status as well as BPV and the study outcomes were examined using multivariable-adjusted Cox proportional hazards models. BPV was analyzed primarily as a continuous standardized variable. Hazard ratios (HRs) and 95% confidence intervals (CIs) were estimated per one-standard-deviation increase in BPV.

Second, potential non-linear associations between BPV and study outcomes were assessed using natural cubic splines with three degrees of freedom, and evidence of non-linearity was evaluated using likelihood ratio tests comparing spline-based models with corresponding linear models (20).

Third, to evaluate the primary study objective, participants were classified according to glycemic status and BPV tertiles to create combined exposure groups. Individuals with normoglycemia and low BPV served as the reference group, and HRs with corresponding 95% CIs were estimated for each joint exposure category.

To facilitate clinical interpretation, predicted 10-year absolute risks were estimated for each glycemia-BPV joint exposure category using the primary confounder-adjusted model. A 10-year prediction horizon was selected because it is clinically relevant and well supported by the study’s median follow-up of 5.1 years.

Fourth, effect modification of glycemic status by BPV was formally evaluated by including multiplicative interaction terms between continuous BPV and glycemic status in pooled models. Statistical significance of interaction was assessed using Wald tests.

Finally, attenuation analyses were performed to determine whether BPV explained part of the excess risk associated with prediabetes and T2D. Associations between glycemic status and the study outcomes were first estimated using multivariable-adjusted Cox models and were then re-estimated after additional adjustment for BPV. Percentage attenuation was calculated using the corresponding log HRs before and after BPV adjustment.

Models were fitted hierarchically. Model 1 adjusted for age, sex, ethnic group, educational attainment, and Townsend deprivation index. Model 2 additionally adjusted for interval mean blood pressure to account for average blood pressure levels. Model 3 was further adjusted for BMI, smoking status, alcohol consumption, and use of blood pressure-lowering and lipid-lowering medication and was considered the primary confounder-adjusted model. Model 4 was additionally adjusted for a history of vascular disease and was treated as a sensitivity model to evaluate whether associations were independent of underlying vascular burden.

### 2.7 Sensitivity Analyses

Several sensitivity analyses were performed to evaluate the robustness of the findings. First, Fine-Gray subdistribution hazard models were fitted for dementia and stroke outcomes to account for the competing risk of death, with associations reported as subdistribution hazard ratios (SHRs) and 95% CIs (21). Second, to reduce the possibility of reverse causation, all primary analyses were repeated after excluding events occurring within the first three years of follow-up. Third, temporal stability of the observed associations was assessed by stratifying follow-up time into 0-5 years and >5 years. Finally, additive interaction was evaluated using the relative excess risk due to interaction (RERI), attributable proportion due to interaction (AP), and synergy index (SI). Positive values of RERI and AP, and SI values greater than 1 were interpreted as evidence of positive additive interaction (22). An additional sensitivity analysis defined SBP variability using the standard deviation (SD) and coefficient of variation (CV) across repeated BP measurements. Associations with mortality, dementia, and stroke were then re-evaluated using interaction models with baseline HbA1c categorized as <48 versus ≥48 mmol/mol (equivalent to <6.5% versus ≥6.5%), as well as baseline HbA1c modelled as a continuous variable. Finally, to evaluate whether the joint associations varied by the severity of prediabetes, we further divided prediabetes into HbA1c 5.7-5.9% (early prediabetes) and HbA1c 6.0-6.4% (late prediabetes) (23).

Benjamini-Hochberg false discovery rate (FDR) correction was applied within each analysis, outcome, and model. For the continuous BPV analyses, the correction set comprised the eight BPV measures representing absolute and directional variability of SBP, DBP, MAP, and PP. All analyses were conducted using R version 4.5.1. Statistical significance was defined as a two-sided p-value <0.05.

## Results

A total of 57,611 participants were included in the analysis and were followed for a median of 5.1 years (IQR, 3.3-9.5 years) from the start-of-follow-up date (mean age 55.4, 52% female). Baseline characteristics stratified by glycemic status are presented in *Table 1*. Compared with participants with normoglycemia (N = 50,744), those with prediabetes (N = 5,397) and T2D (N = 1,470) were older and had a higher prevalence of adverse cardiometabolic profiles. Individuals with T2D had higher BMI, were more likely to have a history of vascular disease and had a higher use of blood pressure-lowering and lipid-lowering medications. They also exhibited less favorable lifestyle characteristics, including higher proportions of current smoking and lower levels of physical activity. Mean SBP, MAP, and PP were higher in participants with prediabetes and T2D than in those with normoglycemia, whereas differences in DBP were modest.

**Table 1.** Baseline characteristics by glycemic status.

| Characteristic | Normoglycemia<br>(N = 50,744) | Prediabetes<br>(N = 5,397) | Type 2 Diabetes<br>(N = 1,470) | P-value |
| --- | --- | --- | --- | --- |
| **Age, years** | 54.9 (7.6) | 58.9 (6.5) | 58.4 (6.9) | <0.001 |
| **Sex, n (%)** |  |  |  | <0.001 |
| Female | 26,650 (52.5%) | 2,795 (51.8%) | 477 (32.4%) |  |
| Male | 24,094 (47.5%) | 2,602 (48.2%) | 993 (67.6%) |  |
| **Ethnic group, n (%)** |  |  |  | <0.001 |
| White | 49,449 (97.4%) | 5,076 (94.1%) | 1,366 (92.9%) |  |
| Other / do not know / prefer not to answer | 1,295 (2.6%) | 321 (5.9%) | 104 (7.1%) |  |
| **Townsend deprivation index, n (%)** |  |  |  | <0.001 |
| Quintile 1 | 10,164 (20.0%) | 1,102 (20.4%) | 247 (16.8%) |  |
| Quintile 2 | 10,220 (20.2%) | 1,046 (19.4%) | 247 (16.8%) |  |
| Quintile 3 | 10,158 (20.0%) | 1,048 (19.4%) | 307 (20.9%) |  |
| Quintile 4 | 10,143 (20.0%) | 1,083 (20.1%) | 286 (19.5%) |  |
| Quintile 5 | 10,016 (19.8%) | 1,114 (20.7%) | 382 (26.0%) |  |
| **Education, n (%)** |  |  |  | <0.001 |
| College / University degree | 23,045 (45.4%) | 2,199 (40.7%) | 527 (35.9%) |  |
| Other | 27,699 (54.6%) | 3,198 (59.3%) | 943 (64.1%) |  |
| **Body mass index, kg/m<sup>2</sup>** | 26.3 (4.0) | 27.8 (4.6) | 30.6 (5.3) | <0.001 |
| **HbA1c, mmol/mol** | 34.1 (32.0, 36.1) | 40.2 (39.5, 41.4) | 50.4 (43.0, 58.3) | <0.001 |
| **Mean arterial pressure, mmHg** | 99.3 (11.6) | 101.6 (11.4) | 100.8 (10.4) | <0.001 |
| **Mean systolic blood pressure, mmHg** | 135.1 (17.8) | 139.5 (17.5) | 139.6 (16.4) | <0.001 |
| **Mean diastolic blood pressure, mmHg** | 81.4 (9.8) | 82.7 (9.9) | 81.4 (9.4) | <0.001 |
| **Mean pulse pressure, mmHg** | 53.7 (12.5) | 56.8 (13.1) | 58.3 (13.5) | <0.001 |
| **Number of contributing blood pressure assessments, n (%)** |  |  |  | 0.002 |
| 2 assessments | 39,914 (78.7%) | 4,345 (80.5%) | 1,195 (81.3%) |  |
| 3 assessments | 10,155 (20.0%) | 1,012 (18.8%) | 260 (17.7%) |  |
| 4 assessments | 675 (1.3%) | 40 (0.7%) | 15 (1.0%) |  |
| **LDL cholesterol, mmol/L** | 3.6 (0.8) | 3.7 (0.9) | 2.8 (0.8) | <0.001 |
| **HDL cholesterol, mmol/L** | 1.5 (0.4) | 1.4 (0.4) | 1.2 (0.3) | <0.001 |
| **Triglycerides, mmol/L** | 1.4 (1.0, 2.0) | 1.6 (1.2, 2.3) | 1.8 (1.2, 2.6) | <0.001 |
| **Physical activity (MET-min/week)** | 1,733.5 (826.0, 3,293.0) | 1,675.0 (810.0, 3,292.0) | 1,404.0 (577.5, 3,048.5) | <0.001 |
| **Smoking status, n (%)** |  |  |  | <0.001 |
| Never | 30,986 (61.1%) | 2,990 (55.4%) | 682 (46.4%) |  |
| Previous | 16,589 (32.7%) | 1,919 (35.6%) | 668 (45.5%) |  |
| Current | 3,053 (6.0%) | 480 (8.9%) | 112 (7.6%) |  |
| **Alcohol status, n (%)** |  |  |  | <0.001 |
| Daily drinker | 26,084 (51.4%) | 2,516 (46.6%) | 559 (38.0%) |  |
| Moderate drinker | 13,097 (25.8%) | 1,349 (25.0%) | 397 (27.0%) |  |
| Social drinker | 9,249 (18.2%) | 1,192 (22.1%) | 401 (27.3%) |  |
| Non-drinker | 2,292 (4.5%) | 337 (6.2%) | 110 (7.5%) |  |
| **Lipid-lowering medication, n (%)** | 6,609 (13.0%) | 1,440 (26.7%) | 1,092 (74.3%) | <0.001 |
| **Blood pressure-lowering medication, n (%)** | 6,497 (12.8%) | 1,335 (24.7%) | 814 (55.4%) | <0.001 |
| **Glucose-lowering medication, n (%)** | 0 (0.0%) | 0 (0.0%) | 1,017 (69.2%) | <0.001 |
| **Vascular disease history, n (%)** | 5,438 (10.7%) | 874 (16.2%) | 470 (32.0%) | <0.001 |

**Abbreviations:** HbA1c, glycated hemoglobin; SBP, systolic blood pressure; DBP, diastolic blood pressure; MAP, mean arterial pressure; PP, pulse pressure; LDL, low-density lipoprotein; HDL, high-density lipoprotein; MET, metabolic equivalent of task.
**Notes:** Baseline assessments were obtained at Visit 0 (Figure 1). BPV was not available at baseline because its calculation required at least two blood pressure assessments.
Data are presented as mean (SD), median (IQR), or n (%), as appropriate. HbA1c, triglycerides, and physical activity are presented as median (IQR); all other continuous variables are presented as mean (SD).
Descriptive statistics were calculated using available data, and percentages for categorical variables were calculated among participants with non-missing data. Missing data were observed for physical activity (n = 9,220; 16.0%), HDL cholesterol (n = 7,256; 12.6%), LDL cholesterol (n = 2,609; 4.5%), triglycerides (n = 2,527; 4.4%), and other variables (<1% each). Consequently, category counts may not sum to group totals.

Consistent with our previous findings (5), associations with all-cause mortality, dementia, and stroke were generally stronger among participants with T2D relative to normoglycemia, than among those with prediabetes, where risks were not always consistently higher than those for normoglycemia *(Supplementary Table S1)*.

### 3.1 Independent associations of BPV with outcomes

In primary confounder-adjusted analyses (Model 3), higher absolute BPV was associated with increased risks of all-cause mortality, dementia, and stroke independent of glycemic status and other confounders *(Figure 2) (Supplementary Table S1)*.

**Figure 2.**
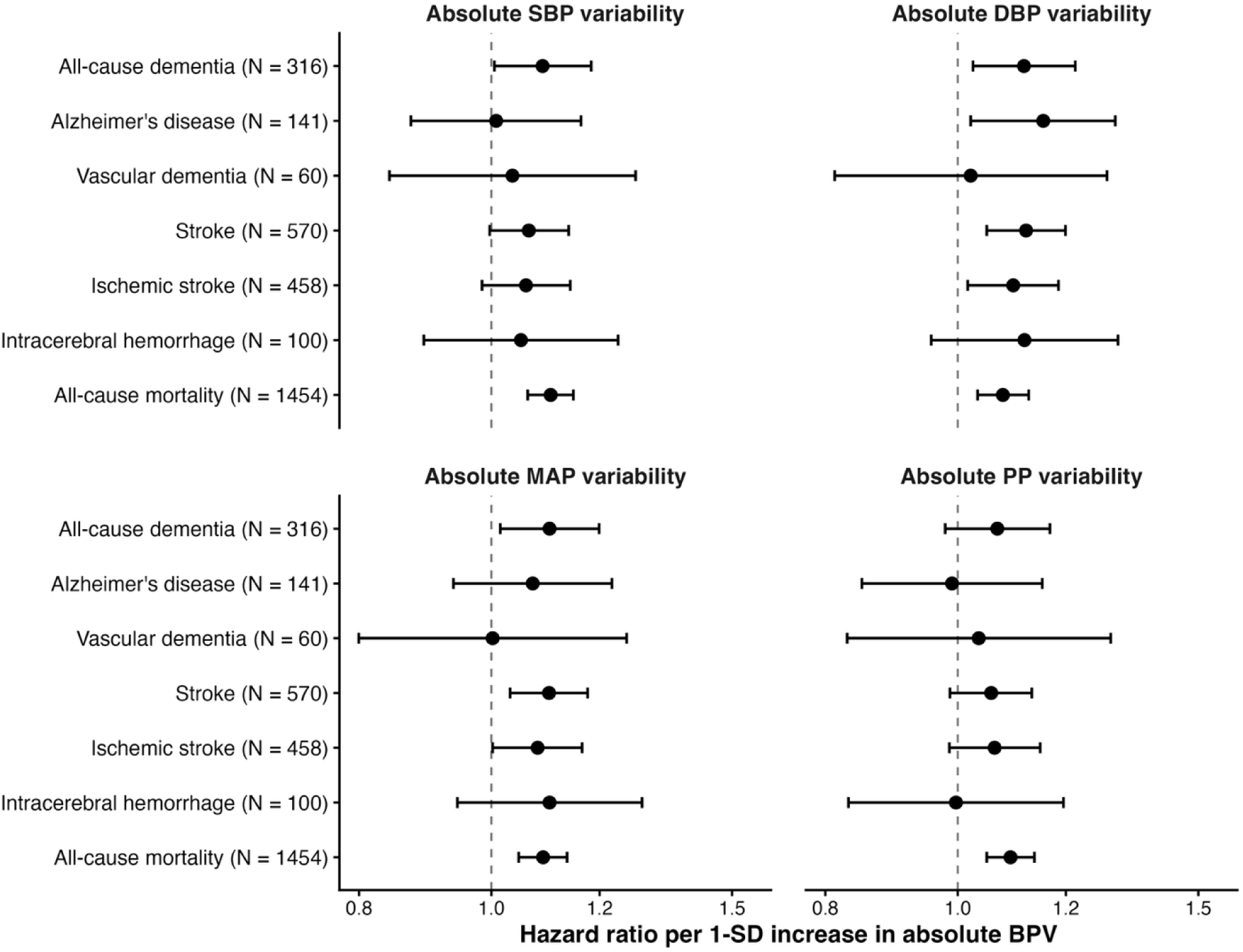
Independent associations of continuous visit-to-visit blood pressure variability with dementia, stroke, and mortality **Abbreviations**: AD, Alzheimer’s disease; BPV, blood pressure variability; CI, confidence interval; DBP, diastolic blood pressure; HR, hazard ratio; ICH, intracerebral hemorrhage; MAP, mean arterial pressure; PP, pulse pressure; SBP, systolic blood pressure; SD, standard deviation; VaD, vascular dementia. **Notes**: HRs and 95% CIs for dementia, stroke, and mortality per one SD higher BPV. Estimates were derived from Model 3 and were adjusted for glycemic status, age, sex, ethnic group, educational attainment, Townsend deprivation index, interval mean blood pressure, body mass index, smoking status, alcohol consumption, and blood pressure- and lipid-lowering medications. Red points indicate nominal statistical significance (P < 0.05); grey points indicate non-significant associations. **Alt text:** Four-panel forest plot showing adjusted hazard ratios and 95% confidence intervals for associations of absolute systolic, diastolic, mean arterial, and pulse pressure variability with dementia, stroke, and mortality. Estimates are presented per standard deviation increase in variability across seven clinical outcomes. Higher absolute variability measures were generally associated with increased mortality risk, while diastolic and mean arterial pressure variability were associated with overall stroke. Associations with dementia were less consistent. Vertical dashed lines indicate a hazard ratio of one, representing no association.

For all-cause dementia, higher BPV was associated with elevated risk. HRs per one-SD increase were 1.09 (95% CI 1.01-1.18) for SBP variability, 1.12 (95% CI 1.03-1.22) for DBP variability, and 1.10 (95% CI 1.02-1.20) for MAP variability; however, none remained significant after FDR correction. A nominal association was also observed between DBP variability and AD (HR 1.16, 95% CI 1.02-1.30; FDR P=0.165). In contrast, higher DBP and MAP variability remained significantly associated with overall stroke after FDR correction, with HRs of 1.12 (95% CI 1.05-1.20; FDR P=0.005) and 1.10 (95% CI 1.03-1.18; FDR P=0.014), respectively. Similar positive associations were observed for IS, but they did not remain significant after FDR correction.

For all-cause mortality, all absolute BPV measures were significantly associated with higher risk after FDR correction. Each 1-SD increase in SBP variability was associated with an 11% higher risk of mortality (HR 1.11, 95% CI 1.06-1.15; FDR P<0.001). Corresponding associations were observed for DBP variability (HR 1.08, 95% CI 1.03-1.13; FDR P=0.001), MAP variability (HR 1.09, 95% CI 1.05-1.14; FDR P<0.001), and PP variability (HR 1.09, 95% CI 1.05-1.14; FDR P<0.001). Further adjustment for vascular history modestly attenuated effect estimates, but the overall pattern of associations remained unchanged.

### 3.2 Non-linear Associations of BPV With Outcomes

Natural cubic spline analyses provided little evidence of substantial non-linearity in the associations between absolute BPV measures and study outcomes (*Supplementary Table S2*). Associations of absolute SBP, DBP, MAP, and PP variability with mortality, dementia, and stroke outcomes were generally consistent with a linear relationship (all FDR-adjusted P-values for non-linearity >0.05). Significant departures from linearity were observed only for selected directional BPV measures to all-cause mortality, including directional SBP variability (FDR P=0.01), MAP variability (FDR P=0.03), and PP variability (FDR P=0.01). Directional DBP variability showed borderline evidence of non-linearity for overall stroke (nominal P=0.01; FDR P=0.05). No FDR-significant departures from linearity were observed for dementia, AD, VaD, IS, and ICH.

### 3.3 Joint associations of glycemic status and BPV

Across all absolute BPV measures, marked variation in risk was observed across the combined glycemia-BPV profiles *(Figure 3) (Supplementary Table S3)*. For all-cause dementia, nominally elevated risks were observed mainly among participants with T2D. Compared with normoglycemia and low BPV, higher risks were observed for T2D combined with high SBP variability (HR 2.46, 95% CI 1.37-4.42), high DBP variability (HR 2.44, 95% CI 1.35-4.43), intermediate DBP variability (HR 2.36, 95% CI 1.10-5.05), high MAP variability (HR 2.07, 95% CI 1.11-3.87), low MAP variability (HR 3.00, 95% CI 1.41-6.38), intermediate PP variability (HR 2.10, 95% CI 1.03-4.29), and low PP variability (HR 2.26, 95% CI 1.07-4.79). However, none remained significant after FDR correction. Findings for AD and VaD were likewise imprecise and FDR nonsignificant because of the limited number of events.

**Figure 3.**
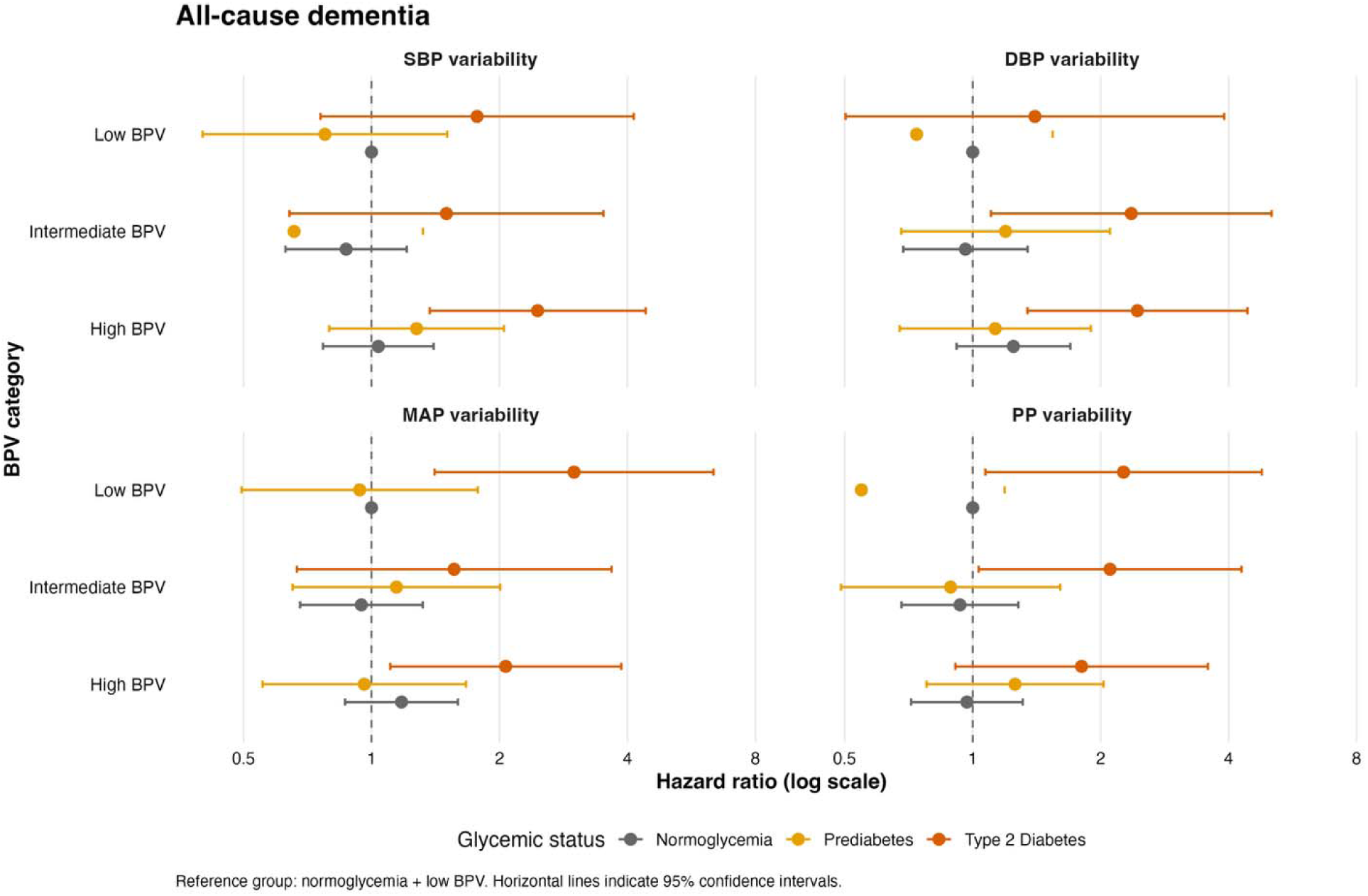
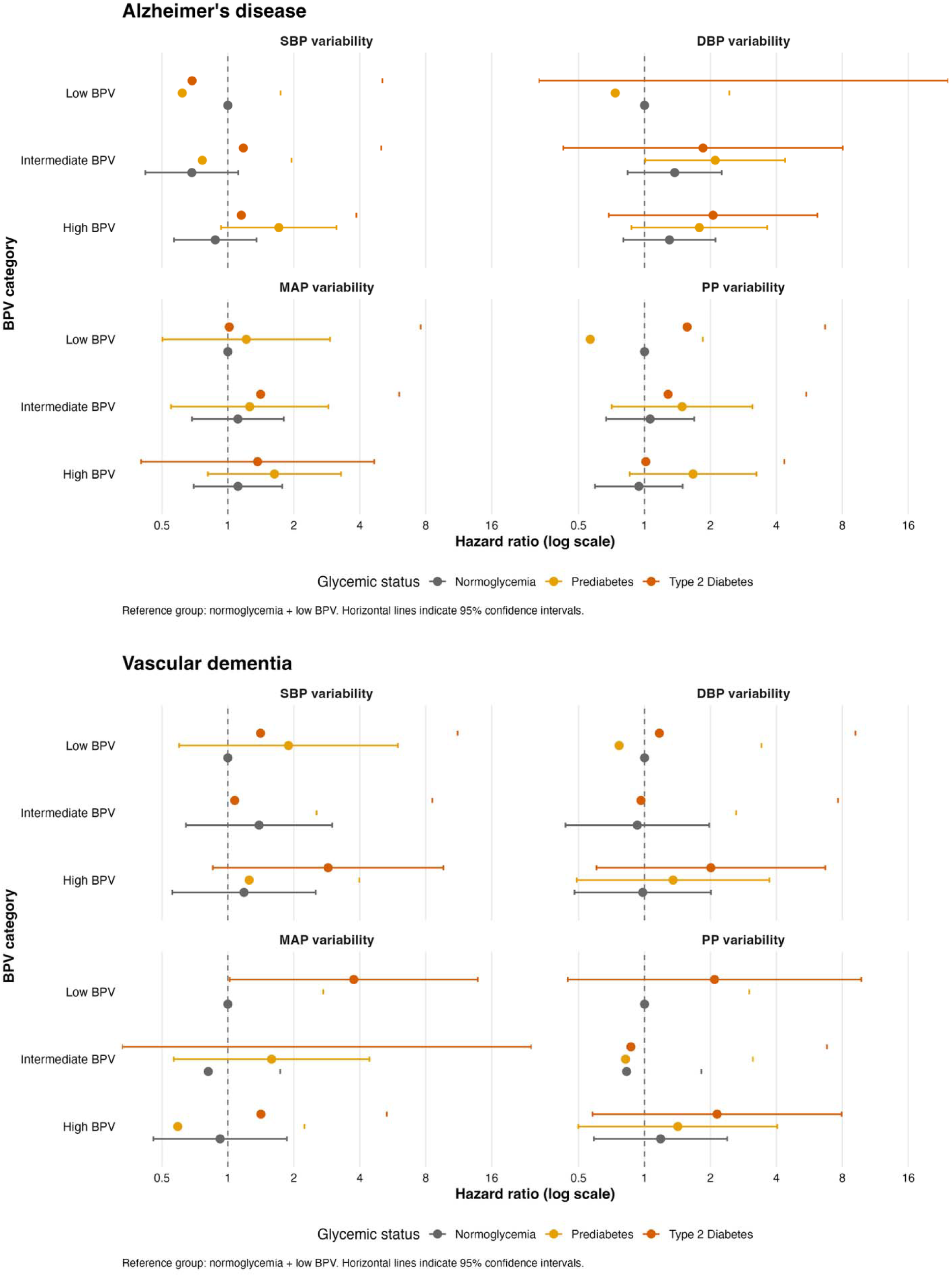
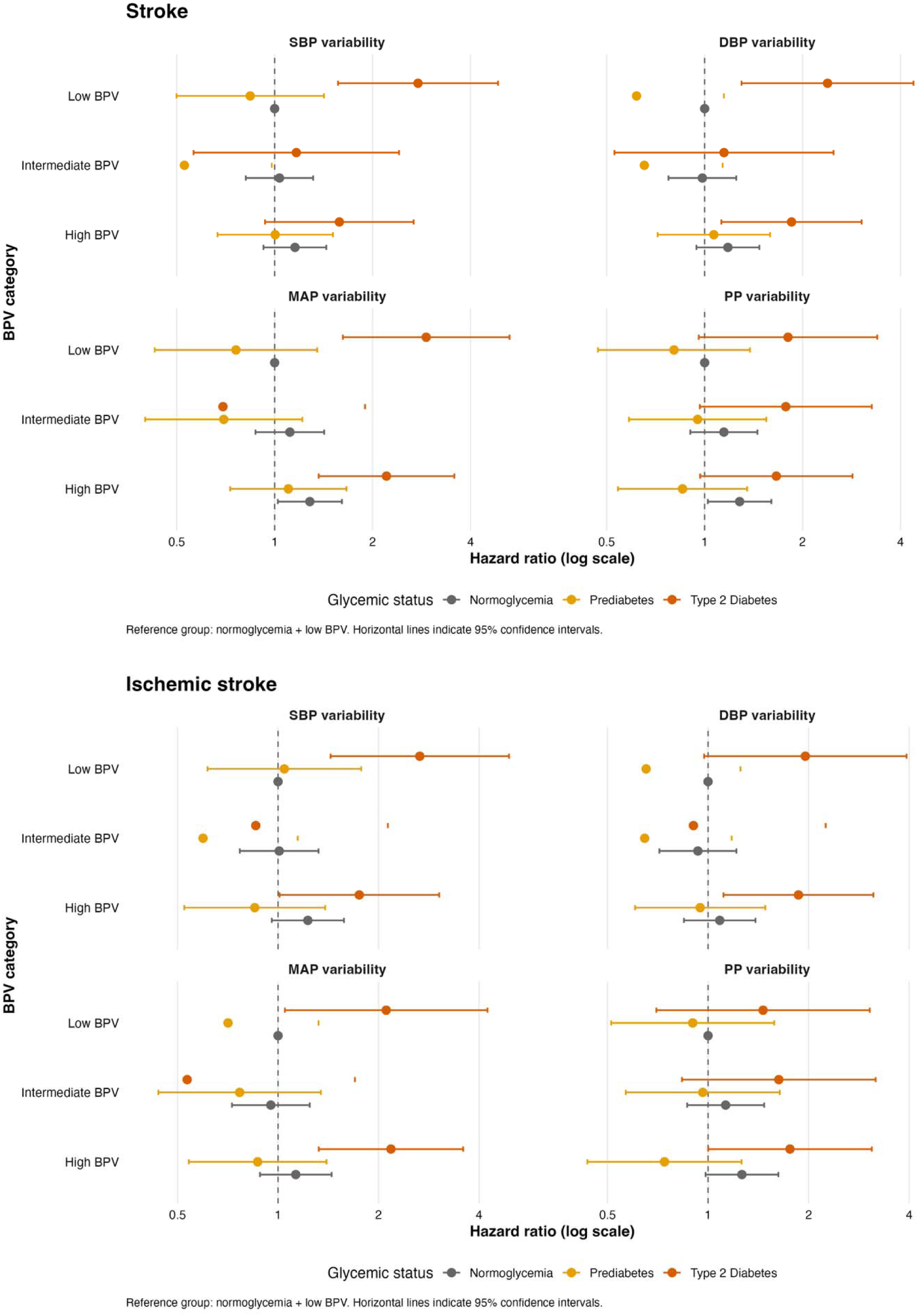
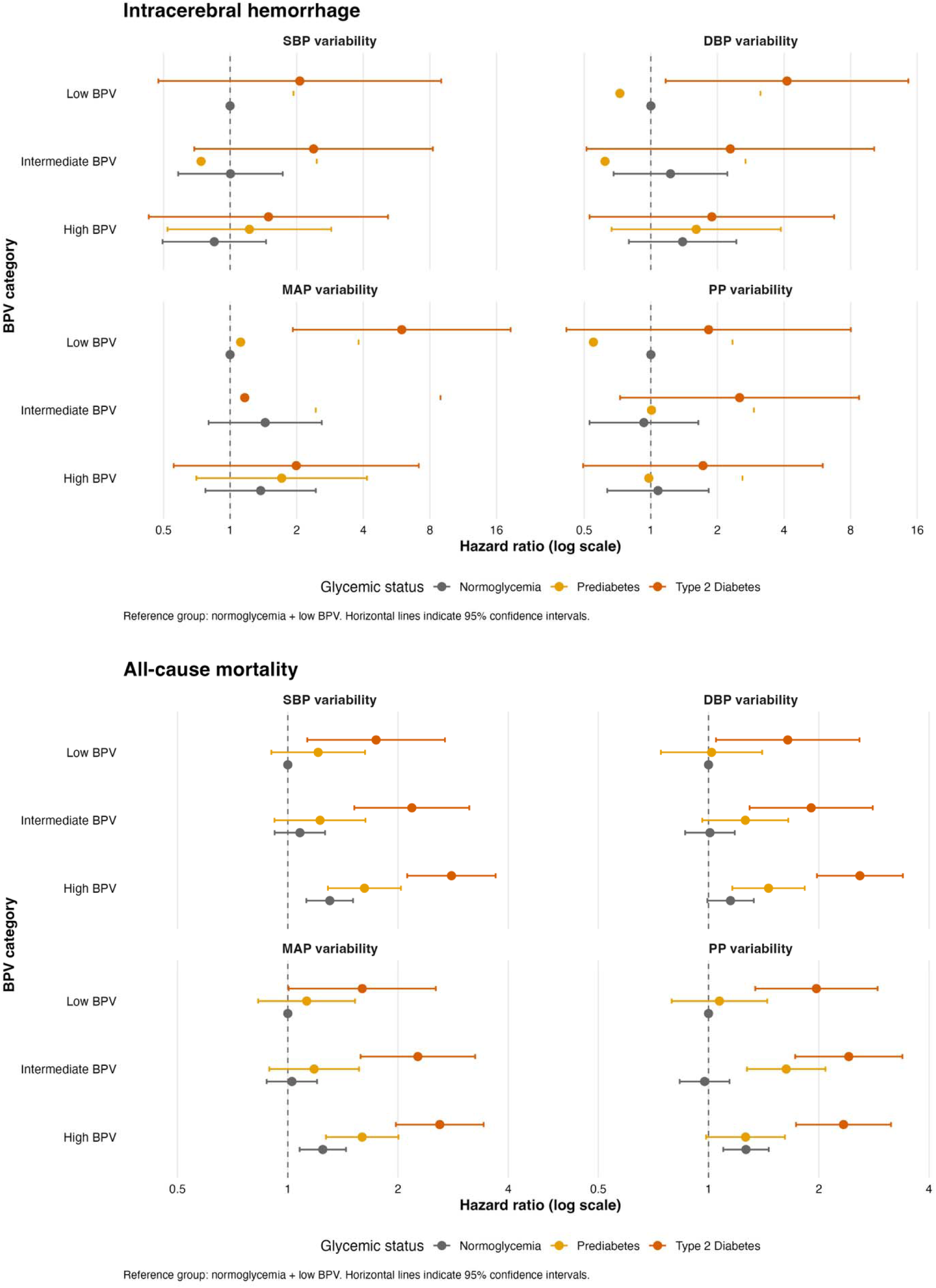
Joint associations of glycemic status and blood pressure variability with risk of dementia, stroke, and all-cause mortality **Abbreviations**: BPV, blood pressure variability; CI, confidence interval; DBP, diastolic blood pressure; HR, hazard ratio; MAP, mean arterial pressure; PP, pulse pressure; SBP, systolic blood pressure; T2D, type 2 diabetes. **Notes**: Forest plots show multivariable-adjusted hazard ratios and 95% confidence intervals for dementia, stroke, and mortality according to combined glycemic status and absolute BPV categories. Estimates were derived from Model 3, adjusted for age, sex, ethnic group, educational attainment, Townsend deprivation index, interval mean blood pressure, body mass index, smoking status, alcohol consumption, and blood pressure- and lipid-lowering medications. The reference group was participants with normoglycemia and low BPV. Horizontal lines indicate 95% CIs. **Alt text:** Seven sets of forest plots showing adjusted hazard ratios and 95% confidence intervals for the joint associations of glycemic status and absolute blood pressure variability with dementia, stroke, and all-cause mortality. Each set includes systolic, diastolic, mean arterial, and pulse pressure variability, categorized as low, intermediate, or high. The reference group is normoglycemia with low variability. Higher risks were generally observed among participants with type 2 diabetes, particularly for all-cause mortality, while associations with dementia and stroke were less consistent.

For stroke, T2D combined with high MAP variability (HR 2.21, 95% CI 1.37-3.57), low MAP variability (HR 2.92, 95% CI 1.62-5.27), and low SBP variability (HR 2.76, 95% CI 1.57-4.86) remained significantly associated with higher risk after FDR correction. Additional nominal associations were observed for T2D combined with high or low DBP variability, whereas prediabetes with intermediate SBP variability was nominally associated with lower risk. For IS, several T2D-BPV combinations showed elevated risks, with HRs ranging from 1.75 to 2.66, but none remained significant after FDR correction, whereas estimates for ICH were less precise because of the smaller number of events.

The strongest and most consistent joint associations were observed for all-cause mortality. Compared with participants with normoglycemia and low BPV, those with T2D and high absolute SBP, DBP, MAP, and PP variability had HRs of 2.80 (95% CI 2.12-3.69), 2.59 (95% CI 1.98-3.39), 2.60 (95% CI 1.97-3.43), and 2.34 (95% CI 1.73-3.15), respectively; all remained significant after FDR correction. Elevated mortality risks were also observed among participants with prediabetes and high SBP, DBP, and MAP variability, with HRs of 1.62 (95% CI 1.29-2.04), 1.46 (95% CI 1.16-1.83), and 1.60 (95% CI 1.27-2.01), respectively. Prediabetes combined with intermediate, rather than high, PP variability was also associated with higher mortality (HR 1.63, 95% CI 1.27-2.09).

Directional BPV analyses showed broadly similar patterns, with the highest risks generally observed among participants with T2D combined with adverse BPV profiles. Additional adjustment for vascular history (Model 4) modestly attenuated effect estimates but did not materially alter the overall pattern of associations.

### 3.4 Predicted 10-year absolute risk according to glycemic status and BPV

Predicted 10-year absolute risks were generally highest among participants with T2D, although gradients across BPV categories were not consistent and estimates were imprecise *(Figure 4) (Supplementary Table S4)*.

**Figure 4.**
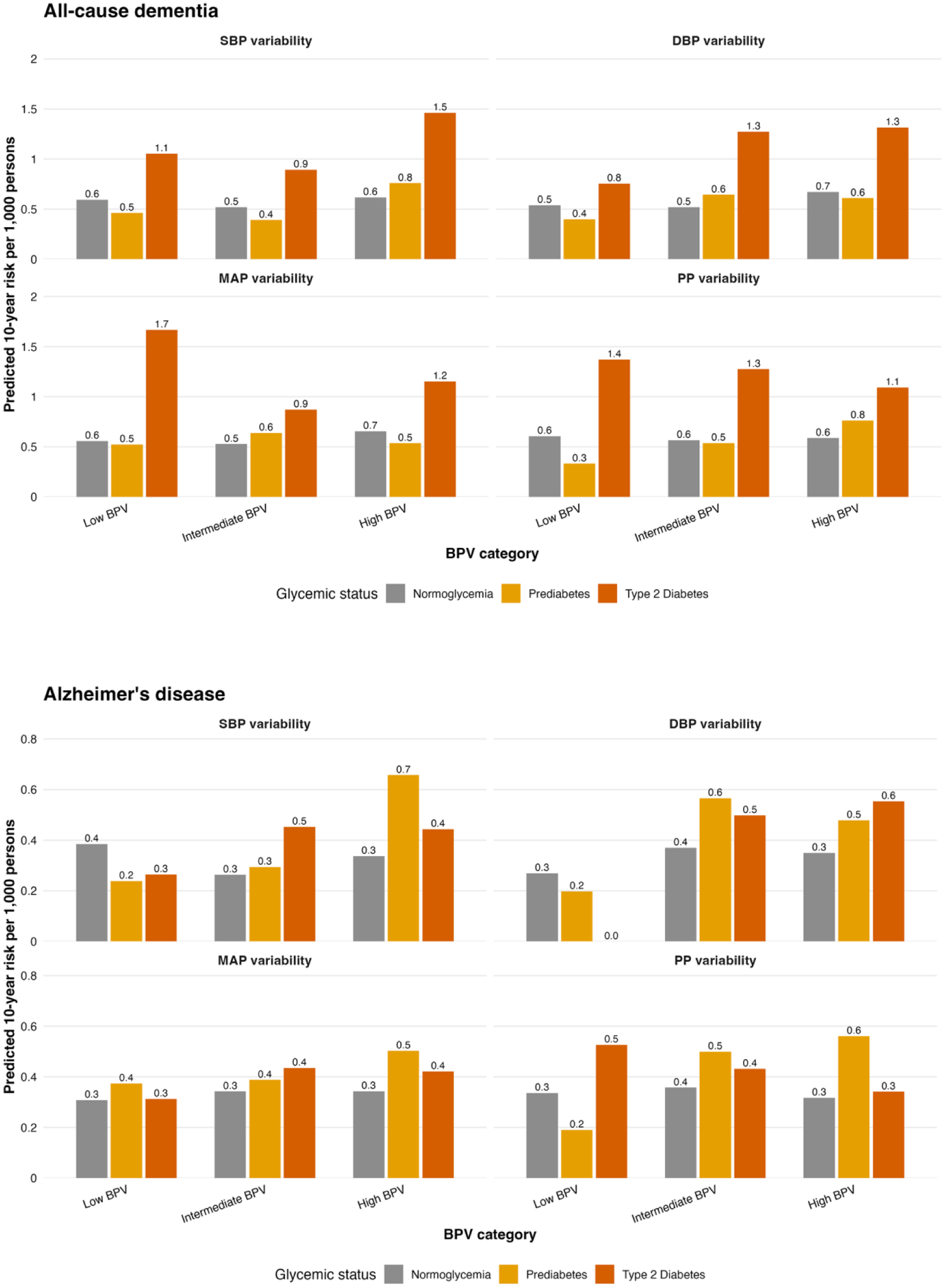
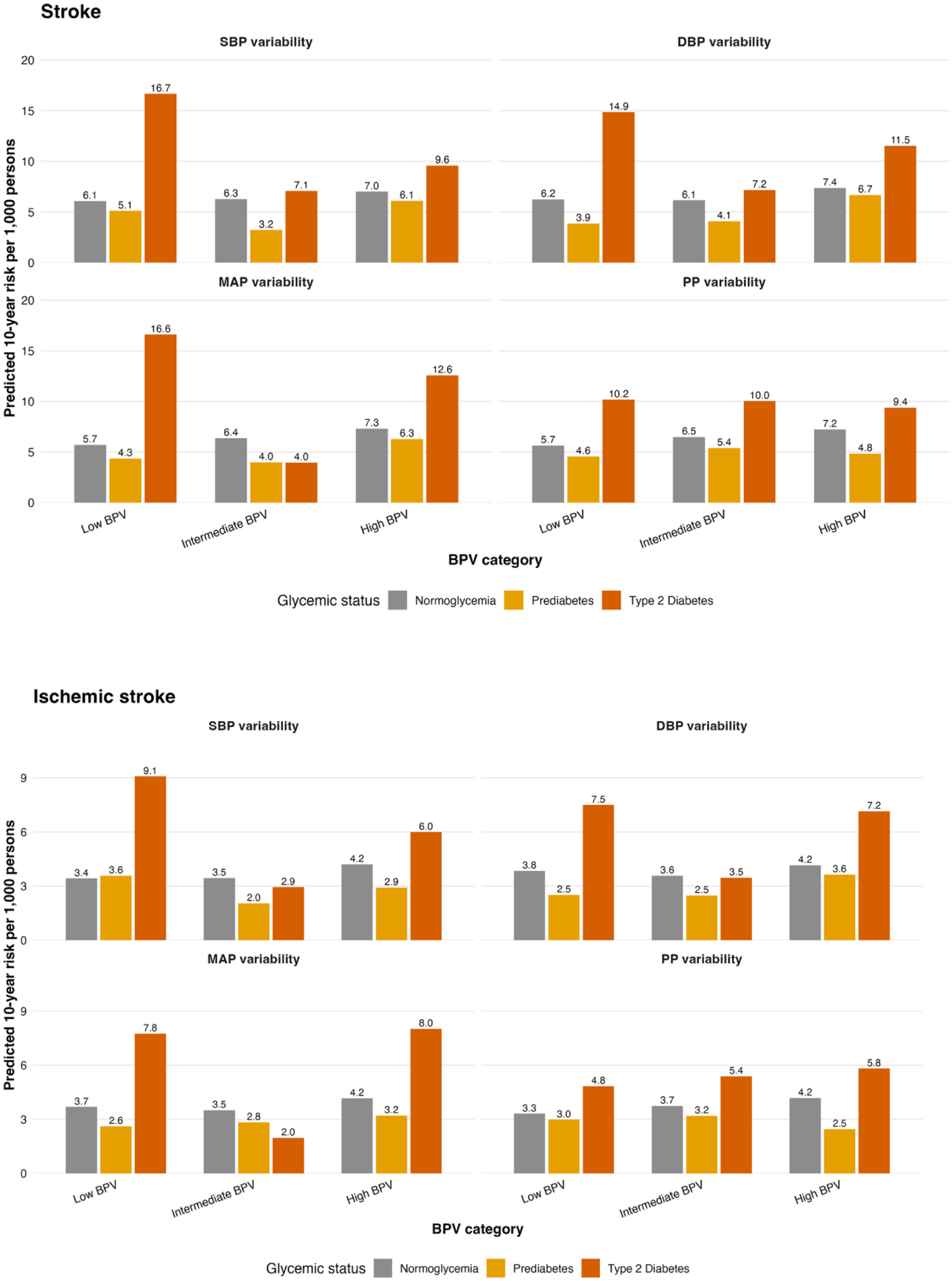
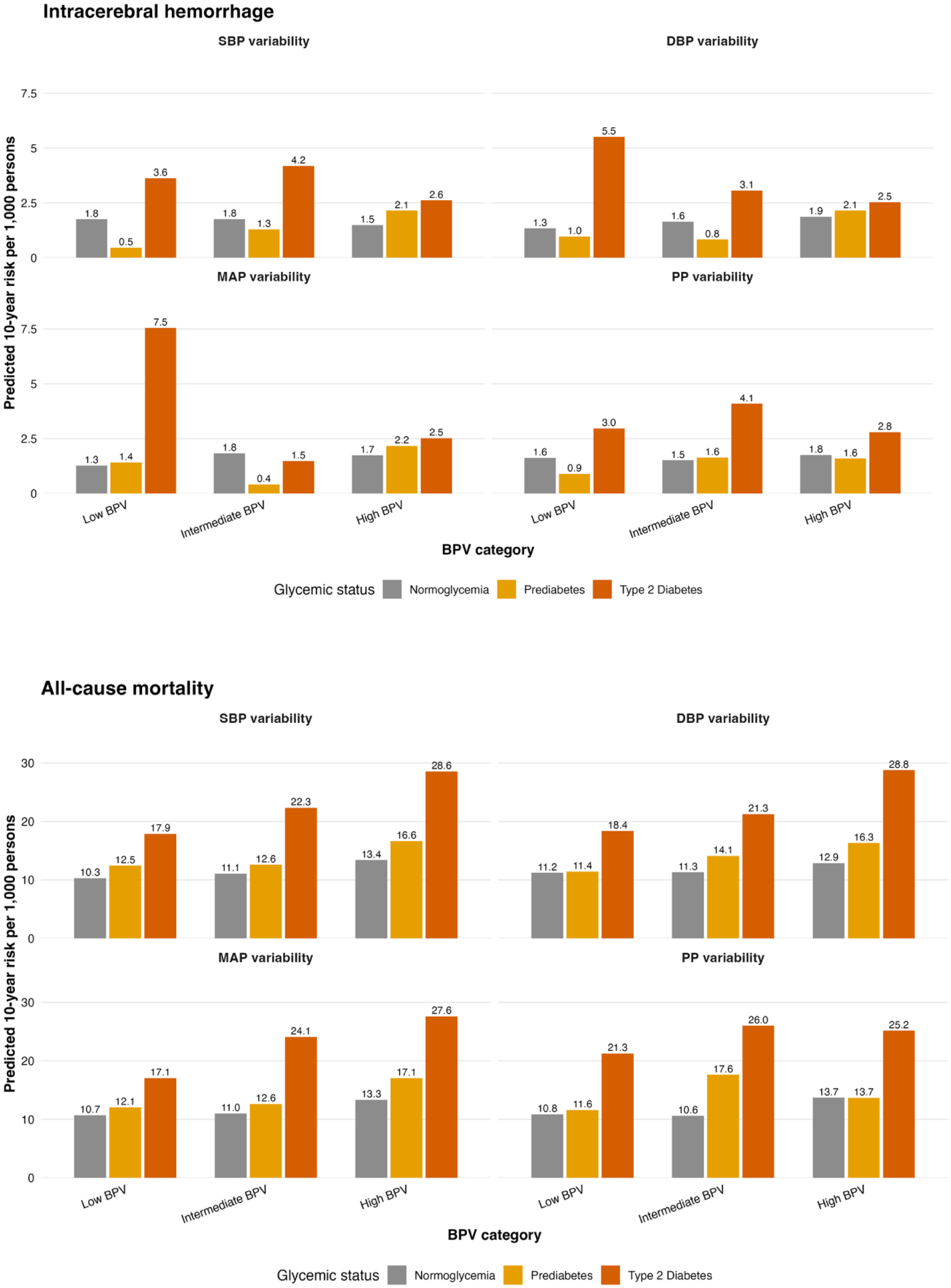
Predicted 10-year absolute risks according to combined glycemic status and blood pressure variability categories **Abbreviations:** BPV, blood pressure variability; DBP, diastolic blood pressure; MAP, mean arterial pressure; PP, pulse pressure; SBP, systolic blood pressure; T2D, type 2 diabetes. **Notes:** Bars represent the model-predicted 10-year absolute risk per 1,000 individuals according to combined glycemic status and BPV categories. Predicted risks were derived from Model 3, adjusted for age, sex, ethnic group, educational attainment, Townsend deprivation index, interval mean blood pressure, body mass index, smoking status, alcohol consumption, and blood pressure- and lipid-lowering medications. SBP, DBP, MAP, and PP variability were each categorized into tertiles (low, intermediate, and high). **Alt text:** Six sets of grouped bar charts showing model-predicted 10-year absolute risks per 1,000 individuals for dementia, stroke, and all-cause mortality across glycemic status and blood pressure variability categories. Each set presents systolic, diastolic, mean arterial, and pulse pressure variability, categorized as low, intermediate, or high. Grey, yellow, and orange bars represent normoglycemia, prediabetes, and type 2 diabetes, respectively. Predicted risks were generally highest among participants with type 2 diabetes, particularly for stroke and all-cause mortality. Risk patterns across blood pressure variability categories differed by outcome and glycemic status.

For all-cause dementia, predicted risks were low in absolute terms. For SBP variability, risk increased from 0.6 per 1,000 individuals among participants with normoglycemia and low BPV to 1.5 per 1,000 individuals among those with T2D and high BPV. Corresponding risks increased from 0.5 to 1.3 per 1,000 individuals for DBP variability. For MAP and PP variability, however, the highest predicted risks occurred among participants with T2D and low BPV: 1.7 per 1,000 individuals for MAP and 1.4 per 1,000 individuals for PP, compared with 1.2 and 1.1 per 1,000 individuals, respectively, among individuals with T2D and high BPV. Predicted risks for VaD were not presented because of the limited number of events and instability of the estimates. For stroke, predicted risks were also generally higher among participants with T2D, but they did not increase consistently with BPV category. For SBP variability, predicted risk was 6.1 per 1,000 individuals among participants with normoglycemia and low BPV, 9.6 per 1,000 individuals among those with T2D and high BPV, and 16.7 per 1,000 individuals among those with T2D and low BPV. Corresponding risks for DBP variability were 6.2, 11.5, and 14.9 per 1,000 individuals; for MAP variability, 5.7, 12.6, and 16.6 per 1,000 individuals; and for PP variability, 5.7, 9.4, and 10.2 per 1,000 individuals, respectively. Participants with prediabetes generally exhibited intermediate risks between those with normoglycemia and T2D.

The clearest absolute-risk differences were observed for all-cause mortality. For SBP variability, predicted mortality risk increased from 10.3 per 1,000 individuals among participants with normoglycemia and low BPV to 28.6 per 1,000 individuals among those with T2D and high BPV. Similar increases were observed for DBP variability, from 11.2 to 28.8 per 1,000 individuals, and for MAP variability, from 10.7 to 27.6 per 1,000 individuals. For PP variability, the predicted risk was 10.9 per 1,000 individuals among participants with normoglycemia and low BPV and 25.2 per 1,000 individuals among those with T2D and high BPV, although the highest estimate occurred in the T2D and intermediate-BPV group at 26.0 per 1,000 individuals. Participants with prediabetes and high BPV also exhibited elevated mortality risk, with predicted risks of 16.6, 16.3, 17.1, and 13.7 per 1,000 individuals for SBP, DBP, MAP, and PP variability, respectively.

### 3.5 Effect Modification of BPV by Glycemic Status

There was little evidence that glycemic status modified the associations between BPV and study outcomes *(Supplementary Tables S5-S6)*. Interaction tests were consistently non-significant across absolute and directional BPV measures, including for all-cause mortality (e.g., interaction *P* = 0.968, 0.460, 0.643, and 0.759 for absolute SBP, DBP, MAP, and PP variability, respectively). Most interaction tests for dementia and stroke were also non-significant. A nominal interaction was observed between absolute PP variability and all-cause dementia (P=0.042), but it did not remain significant after FDR correction (FDR P=0.332).

### 3.6 BPV contribution to glycemia-associated risk

Associations between glycemic status and study outcomes were examined before and after adjustment for BPV *(Supplementary Table S7-S8)*. In Model 3, associations of prediabetes with study outcomes were generally weak or null, whereas stronger associations were observed for T2D, particularly for all-cause mortality, all-cause dementia, and overall stroke.

Adjustment for BPV resulted in minimal changes to the estimated associations for prediabetes. For all-cause mortality, the HR associated with prediabetes remained essentially unchanged after adjustment for BPV (e.g., HR 1.20 before versus 1.20 after adjustment for absolute DBP variability), with attenuation estimates generally close to 0%. Similar findings were observed for dementia and stroke outcomes.

For T2D, an additional adjustment for BPV resulted in only modest attenuation of the risk estimates. For example, the HR for all-cause mortality associated with T2D decreased from 2.05 to 2.03 after adjustment for absolute SBP variability and from 2.04 to 2.03 after adjustment for absolute MAP variability, corresponding to approximately 1% attenuation. Likewise, attenuation of T2D-associated risk was modest for all-cause dementia (approximately 0.5%-1.5%) and stroke (approximately 1.1%-2.2%), depending on the BPV metric examined. Results were materially unchanged in additional Model 4.

### 3.7 Sensitivity Analyses

The primary findings were robust across multiple sensitivity analyses. When accounting for the competing risk of death using Fine-Gray subdistribution hazard models, the overall pattern of associations remained largely unchanged *(Supplementary Table S9)*. For example, participants with T2D and high SBP variability had an increased risk of all-cause dementia (SHR 2.09, 95% CI 1.18-3.69), while those with T2D and high MAP variability had an increased risk of stroke (SHR 1.99, 95% CI 1.25-3.19), consistent with the primary analyses.

Excluding events occurring within the first three years of follow-up did not materially alter the observed associations, suggesting that reverse causation was unlikely to explain the findings *(Supplementary Table S10)*.

Time-stratified analyses demonstrated generally consistent associations during both early (0-5 years) and later (>5 years) follow-up periods *(Supplementary Table S11)*. Although some associations appeared stronger during the first five years of follow-up, the direction and overall pattern of results remained stable over time, with elevated risks consistently concentrated among participants with T2D and higher BPV.

Additive interaction analyses provided modest evidence that the combined effects of dysglycemia and elevated BPV exceeded the sum of their individual effects for selected outcomes *(Supplementary Table S12)*. For example, the RERI for T2D and high absolute SBP variability was 0.54 for all-cause mortality, 0.74 for all-cause dementia, and 0.82 for IS with MAP variability, whereas evidence for other outcomes and BPV metrics was less consistent.

In sensitivity analyses using SBP variability defined as the SD or CV across all available visits, there was no evidence of interaction with either HbA1c ≥ 48 mmol/mol or continuous HbA1c for all-cause mortality, dementia, or stroke outcomes. There was little evidence that baseline HbA1c, modelled either as a binary variable (<48 versus ≥48 mmol/mol) or as a continuous measure, modified the associations between SBP variability and mortality, dementia, or stroke outcomes (all interaction P≥0.166; all FDR-adjusted P≥0.206; *Supplementary Table S13-S14*). These findings indicate no detectable HbA1c-related effect modification but do not provide consistent support for the primary findings across alternative BPV definitions.

Sensitivity analyses using a refined four-category glycemic classification (normoglycemia, prediabetes 5.7-5.9%, prediabetes 6.0-6.4%, and T2D) yielded findings broadly similar to the primary joint-group analyses (*Supplementary Table S15*). The clearest associations were observed for mortality: compared with normoglycemia and low BPV, T2D combined with high SBP variability was associated with an almost threefold higher risk (HR 2.79, 95% CI 2.11-3.68), while those with early prediabetes and high SBP variability showed a more modest increase (HR 1.74, 95% CI 1.37-2.21). Associations with dementia and stroke were less consistent across BPV categories.

## Discussion

In this large prospective cohort study, we evaluated the independent, joint, and interactive associations of visit-to-visit BPV and glycemic status with risks of dementia, stroke, and all-cause mortality. Four principal findings emerged. First, higher absolute BPV was independently associated with all-cause mortality after accounting for glycemic status and mean blood pressure. Higher DBP and MAP variability were also associated with overall stroke after correction for multiple testing, whereas associations with dementia and IS were nominal and did not remain significant after FDR correction. Second, regarding glycemic status, T2D was more strongly associated with the outcomes than prediabetes, where risks were not consistently elevated compared with normoglycemia. Third, joint associations between glycemic status and BPV were strongest and most consistent for all-cause mortality, particularly among individuals with T2D. For dementia and stroke, elevated risks were observed across several T2D-BPV combinations, but there was no consistent gradient across BPV categories. Finally, there was little evidence that BPV modified the associations between glycemic status and study outcomes, as no interaction remained significant after FDR correction, and adjustment for BPV minimally attenuated the associations between glycemic status and study outcomes, indicating that BPV did not substantially explain the excess risk associated with dysglycemia.

Our findings are broadly consistent with previous evidence showing that visit-to-visit BPV is associated with increased risks of stroke and all-cause mortality beyond mean blood pressure. Higher BPV has been associated with dementia independent of antihypertensive treatment and with greater risks of cardiovascular events, chronic kidney disease, dementia, and death irrespective of average blood pressure (11, 15). Recent evidence also suggests that BPV and glycemic variability contribute to vascular injury through shared mechanisms, including endothelial dysfunction, oxidative stress, and arterial stiffness, and that their coexistence is associated with a substantially greater risk of cardiovascular morbidity and mortality in individuals with T2D (16, 24, 25). Our results extend this evidence by evaluating several BPV metrics alongside glycemic status. However, the most robust associations were observed for mortality, while findings for dementia and stroke subtypes were less consistent and should be interpreted cautiously.

A key finding was that the joint associations of glycemic status and BPV were strongest and most consistent for all-cause mortality. Participants with T2D generally had the highest relative and predicted absolute risks, although the ordering of BPV categories was less consistent for dementia and stroke. Predicted 10-year mortality risk increased from approximately 1% among participants with normoglycemia and low BPV to nearly 2.5%-2.9% among those with T2D and high BPV. These absolute-risk estimates complement the hazard ratios by illustrating the potential difference in expected risk between clinically relevant subgroups, although their wide confidence intervals warrant cautious interpretation. The relatively low predicted 10-year risk of dementia, including AD, may partly reflect the younger age of the study population, the long latency of dementia development, and the competing risk of death.

Interestingly, the associations differed across BP variability metrics. All four absolute BPV measures were independently associated with all-cause mortality after FDR correction, whereas only DBP and MAP variability remained associated with overall stroke. Nominal associations were observed between SBP, DBP, and MAP variability and all-cause dementia, between DBP variability and AD, and between DBP and MAP variability and IS, but these did not survive multiple-testing correction. Joint analyses similarly showed the clearest and most consistent associations for mortality, while patterns for dementia and stroke varied across BPV measures and categories. Thus, the findings do not support one BPV metric as uniformly superior across all outcomes, although DBP and MAP variability appeared to show the broadest patterns of association.

These differences are biologically plausible because the various BPV measures capture distinct aspects of blood pressure regulation. SBP variability primarily reflects fluctuations in large-artery pressure and arterial stiffness, which have been linked to cerebral small vessel disease and cognitive decline (26, 27). In contrast, DBP variability may have a greater impact on brain health because impaired cerebral autoregulation and microvascular dysfunction in diabetes may render the brain more vulnerable to fluctuations in diastolic pressure (28). MAP variability may reflect fluctuations in tissue perfusion pressure and cumulative hemodynamic stress affecting the cerebral and systemic circulation, while PP variability may reflect fluctuations related to arterial stiffness and vascular aging (29–32). These differences may partly explain the stronger association of PP variability with all-cause mortality than with specific neurological outcomes in our study.

We found little evidence that BPV substantially modified or explained the associations between glycemic status and adverse outcomes. No multiplicative interactions remained significant after multiple-testing correction, and adjustment for BPV attenuated the glycemia-associated hazard ratios by only 0-2%, suggesting that BPV is unlikely to be a major pathway linking dysglycemia to dementia, stroke, or mortality. BPV also appeared to have limited explanatory or effect-modifying value, with alternative SD/CV analyses showing largely null mortality estimates and inverse, rather than consistent, associations with dementia. Instead, BPV appears to provide complementary prognostic information beyond glycemic status. These findings were robust across alternative BPV definitions, HbA1c interaction analyses, natural spline analyses, and multiple sensitivity analyses, including competing-risk, lagged, and time-stratified models.

Several strengths and limitations should be acknowledged. A major strength is the use of repeated routine clinical blood pressure measurements, allowing BPV to be assessed in a real-world setting. The study also evaluated multiple clinically relevant outcomes and examined both the independent and joint associations of BPV and glycemic status, providing a comprehensive assessment of their potential relationships with mortality and neurological outcomes. However, several limitations should be considered. First, the observational design precludes causal inference, and residual confounding cannot be excluded despite extensive adjustment. Second, BPV can be assessed using various methods, and our approach, based on routine clinical measurements performed years apart, may be affected by measurement variability, visit frequency, and treatment changes over time. Variations in BPV definitions and measurement approaches may also account for discrepancies between our findings and those of previous studies reporting stronger associations. Third, the study population had a relatively low incidence of dementia, and the relatively young age of the population and/or duration of follow-up may have limited our ability to detect associations with dementia, particularly for dementia subtypes. Some subgroup analyses, particularly for dementia subtypes and ICH, were also based on relatively small numbers of events and therefore yielded imprecise estimates. Finally, glycemic status was assessed at baseline and may have changed during follow-up, potentially leading to exposure misclassification.

Future studies should assess BPV using standardized and repeated measurements, including alternative BPV metrics, to determine whether the associations are robust across different definitions and measurement approaches. Studies in older populations with longer follow-up and a higher incidence of dementia may also help clarify whether BPV and glycemic status are associated with dementia and its subtypes. Repeated assessment of glycemic status and BPV over time may further help clarify their temporal and potential joint associations with neurological and other clinical outcomes.

Several biological mechanisms may explain the observed associations. Impaired glucose metabolism may promote endothelial dysfunction, inflammation, oxidative stress, vascular remodeling, and microvascular injury(33–40). Within the brain, these processes may contribute to the disruption of cerebral autoregulation, blood-brain barrier dysfunction, and cumulative damage to small cerebral vessels, potentially accelerating neurodegenerative and cerebrovascular processes, and thereby increasing the risks of dementia and stroke (12, 41–43). BPV may contribute to these processes through repeated mechanical stress on the vascular wall and impaired cerebral perfusion. However, the limited attenuation of the associations after accounting for BPV suggests that BPV provides little explanation for the observed associations between glycemic status and neurological outcomes.

These findings may have implications for cardiovascular risk assessment. Current prevention strategies primarily emphasize glycemic control and mean blood pressure, whereas BPV is not routinely considered in clinical practice. In our study, BPV provided prognostic information beyond glycemic status and mean blood pressure, with the most robust associations observed for all-cause mortality and overall stroke. Joint associations were strongest among participants with T2D, particularly for mortality, although patterns across BPV categories were less consistent for dementia and stroke. Together with the differences in predicted absolute risk, these findings suggest that incorporating BPV into longitudinal blood pressure assessment may help identify individuals who warrant closer monitoring and more intensive management of modifiable risk factors. However, further studies are needed to determine whether incorporating BPV into clinical risk prediction improves discrimination, reclassification, or patient outcomes.

In conclusion, the joint associations of dysglycemia and BPV were strongest and most consistent for all-cause mortality, with elevated risks also observed across several stroke and dementia subgroups. Although BPV explained little of the excess risk associated with dysglycemia and there was little evidence that BPV modified glycemia-outcome associations, BPV remained independently associated with all-cause mortality and overall stroke after correction for multiple testing, whereas associations with dementia and stroke subtypes were less consistent. These findings suggest that BPV may provide prognostic information beyond glycemic status and mean blood pressure, although further research is needed to determine its incremental value for clinical risk prediction.

## Funding

This work was supported by the Joint Programme - Neurodegenerative Disease Research (JPND) through ZonMw, the Netherlands Organisation for Health Research and Development (project number 10510062510005), as part of the NeuroSync project. S.H. is supported by a scholarship from the China Scholarship Council (CSC) under Grant No. 202208330062. The study funder was not involved in the study design, the collection, analysis, and interpretation of data, or writing of the report, and did not impose any restrictions regarding the publication of the report.

## Conflicts of Interest

The authors declare that they have no competing interests.

## Author Contributions

F.A. conceived the idea presented. S.H. conducted all statistical analyses under the supervision of F.A., G.J.B., and M.S. J.D. contributed to the analysis plan. S.H. drafted the manuscript, with F.A. providing critical revisions and feedback. Y.Z. helped develop the methodology. All authors have reviewed and approved the final version of the manuscript.

## Availability of data and materials

This research was conducted using the UK Biobank resource under application number 100993. The UK Biobank data are available to bona fide researchers upon application and approval by UK Biobank (www.ukbiobank.ac.uk). Restrictions apply to the availability of these data, which were used under license for the current study and are not publicly available.

## Declaration of Generative AI and AI-assisted technologies in the writing process

During the preparation of this work, the authors used ChatGPT (OpenAI) to assist with language editing, improving clarity, and grammar of the text. The authors critically reviewed, edited, and verified all AI-assisted content and take full responsibility for the accuracy, originality, and integrity of the final manuscript.

## Data Availability

All data produced in the present study are available upon reasonable request to the authors.

## Notes

### Competing Interest Statement

The authors have declared no competing interest.

### Author Declarations

The North West Multi-centre Research Ethics Committee gave ethical approval for the UK Biobank study. All participants provided informed consent, and the present analyses were conducted using data accessed under an approved UK Biobank research application.

